# Avoiding Poor Skull Areas Improves Heating Efficiency in MR-guided Focused Ultrasound Therapy

**DOI:** 10.1101/2025.06.17.25328836

**Authors:** Makoto Kadowaki, Kenji Sugiyama, Mikihiro Shimizu, Takao Nozaki, Kaoru Kikuyama, Akira Okazaki, Muneaki Hashimoto, Tomohiro Yamasaki, Yoshinobu Kamio, Hiroaki Neki, Hiroki Namba, Kazuhiko Kurozumi

**Affiliations:** Center for Clinical Research, Hamamatsu University Hospital, Hamamatsu, Shizuoka, 431-3192, Japan; Department of Neurosurgery, Hamamatsu University School of Medicine, Hamamatsu, Shizuoka, 431-3192, Japan; Department of Neurosurgery, JA Shizuoka Kohseiren Enshu Hospital, Hamamatsu, Shizuoka, 430-0929, Japan; Department of Neurosurgery, Toyoda Eisei Hospital, Iwata, Shizuoka, 438-0838, Japan

**Keywords:** focused ultrasound, heating efficiency, skull density ratio, essential tremor, Parkinson’s disease, thalamotomy

## Abstract

Transcranial magnetic resonance-guided focused ultrasound surgery (MRgFUS) is an effective and safe treatment for drug-resistant symptoms of Parkinson’s disease and essential tremor. In some patients, MRgFUS is difficult to perform because the temperature in the target area does not easily increase. In this study, we evaluated the effectiveness of avoiding the sonication of skull regions with poor conditions and redistributing energy to relatively good-quality regions.

We retrospectively analyzed MRgFUS data at our facility. In some patients, when it was considered difficult to achieve the required temperature in the target area and thermal coagulation after starting treatment, the ultrasound transducer elements that passed through areas with poor conditions, considered as such on the basis of the skull density ratio and skull thickness, were turned off. A linear regression model was created for all ultrasound sonications in all patients, with heating efficiency as the objective variable and various treatment and patient factors, including whether cranial areas with poor conditions were excluded from the sonication field, as explanatory variables.

In total, 213 patients underwent MRgFUS treatment, with 1,891 sonications. Eleven patients had 30 sonications excluding skull areas with poor conditions. The median number of excluded elements was 37.5 (maximum 74, minimum 6). Linear regression analysis showed that excluding cranial regions with poor conditions from the sonication field significantly improved the heating efficiency (*P* = 0.0125, *R*^2^ = 0.769). The regression coefficient was 0.286°C/kJ, which means that for every additional 10,000 J of sonication energy, the temperature increases by 2.86°C. The difficulty of controlling coagulation foci and the number of adverse events were not obviously increased using this method.

When sonication was conducted while avoiding areas with a low skull density ratio and a thick skull, the heating efficiency of MRgFUS significantly improved. Using this approach, even difficult-to-treat cases may be successfully managed.

## Introduction

Transcranial magnetic resonance-guided focused ultrasound surgery (MRgFUS) is an effective and safe treatment for drug-resistant symptoms of Parkinson’s disease and essential tremor^1–3^. MRgFUS can non-invasively coagulate the thalamus and globus pallidus by focusing a large number of ultrasound waves deep in the brain to increase the temperature in the target area. However, in some situations, MRgFUS may have difficulty in increasing the temperature in the target area, making treatment difficult.

The skull density ratio (SDR) is one of the key variables for predicting the therapeutic efficacy of MRgFUS^4–10^. When the SDR is low, the target site temperature is less likely to rise^4,6,7^, more energy is required^5,9,11^, heating efficiency is reduced^12,13^, and adverse events increase in frequency^6^. Recently, however, several retrospective studies^5–7,10^ and one prospective study^14^ have reported that successful MRgFUS treatment can be achieved even with a low SDR, and safety has also been noted^6,15^.

Furthermore, it is known that the temperature in the target area is less likely to increase with each repetition of MRgFUS ultrasound sonication^12,16^. It has been reported that factors such as increasing power^17^, increasing energy^16^, and cumulative temperature increases^12^ reduce heating efficiency. Therefore, performing high-energy sonication at an earlier stage is expected to be more effective^12,16^, and one retrospective study has reported positive results in this regard^18^.

We previously found that the SDR is better at predicting treatment outcomes when calculated after excluding the bilateral temporal regions, which are highly permeable to ultrasound^19^. The skull is known to be thinner in bilateral temporal regions, which have significantly better ultrasound permeability^20^. Taken together, the findings indicate that even in regions with a low SDR, if the skull is thin, clinically effective ultrasound permeability can be achieved. Therefore, we speculate that if we avoid sonicationg skull regions with poor conditions in terms of both the SDR and skull thickness, and redistribute the sonication energy to regions with better conditions, then we can achieve more effective temperature rises.

In MRgFUS treatment using the Exablate4000 device (Insightec, Tirat Carmel, Israel), a focused ultrasound beam is directed at the treatment target from 1,024 ultrasound transducer elements, arranged in a helmet shape, causing the temperature of the tissue to rise and resulting in thermal coagulation. Frontal sinuses or calcified tissues in the sonication path should be set as “No Pass Regions”^21^, and the transducer elements that emit the ultrasound beam that passes through them should be turned off. Insightec further states that, with respect to the number of ultrasound transducer elements, the operator should “ensure minimum values of 700 calculated active elements…to ensure a safe and effective treatment”^21^. However, the basis for this recommendation is not stated, and no published paper provides one either; nevertheless, simulation results show that it becomes difficult to control the size of the coagulation clot when there are fewer than 700 active elements (Insightec Japan, personal communication, January 2025).

In this study, we retrospectively collected cases where the heating efficiency was poor, a sufficient temperature rise was considered difficult after treatment began, and cranial regions with poor skull conditions were excluded from the sonication field, and we evaluated the effects of such exclusion. However, in the overwhelming majority of cases, coagulation foci ultimately formed, and symptomatic improvement was achieved, so it is difficult to statistically evaluate the success or failure of treatment using the formation of a coagulation foci as an endpoint. Therefore, in this study, the relationship between treatment settings and temperature rise was evaluated using the heating efficiency of each sonication as a surrogate endpoint.

## Materials and methods

### Patients

Patients treated at our institution between April 2019 and December 2024 were included in this study. Informed consent was obtained in writing from patients with an SDR_mean_ of 0.40 or less after thoroughly explaining the possibility of treatment failure and complications. Because this was a retrospective study, patient consent was waived, and patients who refused to participate in the study (i.e., who opted out) were excluded.

### MRgFUS procedures

The treatment method was described in detail in a previous paper.^22^ In summary, sonication was performed with the Exablate 4000 (Insightec) system, targeting the ventral intermediate nucleus of the thalamus. The coordinates of the target were tentatively set according to the atlas and then modified on the basis of magnetic resonance imaging. First, sonication was performed with a low-energy dose, and the temperature was increased to approximately 43–47°C. Misalignment between the target of sonication and the site of the temperature increase was then corrected by magnetic resonance thermometry. Next, the energy dose was increased to 48–50°C, and it was confirmed that sonication improved symptoms and had no side effects. Finally, sonication was performed with a higher energy dose to raise the temperature to 53–60°C, and the target area was thermally coagulated. Once the intended temperature rise was achieved, magnetic resonance imaging was performed to confirm that coagulation foci had been created in the target area. Additional sonication was performed if needed.

In cases where a temperature rise above 53°C was difficult, possible temperature elevations were repeated. Treatment was completed when sufficient improvement of symptoms and the formation of T2 lesions were observed. The procedure was discontinued when it was judged that it would be difficult to continue treatment because of intraoperative symptoms such as headache, or when the temperature rise leading to thermal coagulation could not be achieved.

### Exclusion of skull areas with poor conditions

In cases where a sufficient temperature rise was deemed unlikely after treatment had begun, skull regions with poor conditions were excluded from the sonication field (Fig. 1). The skull conditions for each of the 1,024 ultrasound elements can be displayed on the Exablate 4000 treatment console, where elements can be turned off if necessary. Elements that were considered to have poor conditions overall were turned off on the basis of overlapping areas with a low SDR and thick skull. The total number of elements was set to be at least 850, ensuring a sufficient margin relative to the minimum of 700 elements stipulated by the treatment guidelines.

**Figure 1.**
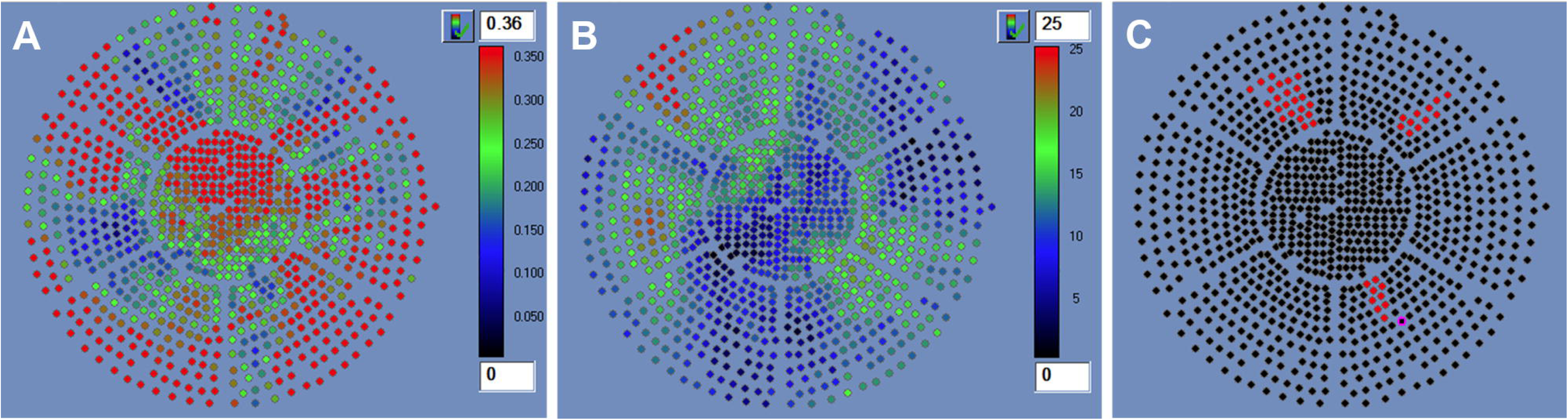
An example of element exclusion. (**A**) Skull density ratio. (**B**) Skull thickness (mm). (**C**) Excluded ultrasound transducer elements. A total of 43 elements with a low skull density ratio and thick skull were excluded.

### Treatment variables

After treatment, by using the export function of the Exablate 4000 device, the number of elements used in the treatment, the sonication energy (J), the sonication power (W), and the maximum average temperature reached (°C) were obtained. To derive the heating efficiency, 37°C was subtracted from the maximum average temperature, and the result was divided by the effective energy used^12,16^.

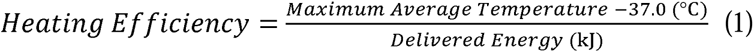

As an indicator of cumulative temperature rise, we used the subtotal of the maximum average temperature reached during the previous sonication minus 37°C^12^. To visualize the treatment variables, we drew a histogram of heating efficiency and the number of sonication elements. We also visualized the relationship between sonication energy and temperature rise for cases that required more than 35,000 J of energy (i.e., cases that required considerable energy for a temperature rise and cases that were difficult to treat), including cases where areas of poor sonication were excluded.

### Statistics

To summarize the data, we calculated the mean and standard deviation for normally distributed continuous variables, the median and interquartile range for non-normally distributed continuous variables, and percentages for binary variables. A Q–Q plot was used to evaluate the normality of the data distribution. To test for differences between the groups, the chi-squared test was used for binary variables, Student’s t-test for continuous variables following a normal distribution, and the Mann–Whitney U test for continuous variables not following a normal distribution. To test for correlations, Spearman’s correlation coefficient was used. To evaluate the impact of treatment factors on heating efficiency, we constructed a linear regression model to predict the heating efficiency of individual sonications. A linear regression model with a robust estimation method was used to take patient clusters into account. The explanatory variables were basic patient background information, including age, gender, diagnosis, and SDR, as well as treatment factors that have been reported in previous studies to be related to heating efficiency, including sonication power, sonication energy, the number of ultrasound elements used, the cumulative number of sonications, and the cumulative temperature rise. We set the significance level at *P* = 0.05. All statistical analyses were performed with R software (The R Foundation for Statistical Computing, Vienna, Austria). For linear regression using a robust estimation and considering patient clusters, the lmrobust function of the estimatr package was used.

### Ethical considerations

This study was approved by Ethics Committee of Hamamatsu University School of Medicine (Approval No. 23-108).

## Results

In total, 213 MRgFUS treatments were performed at our facility, including 1,891 sonications. Of these cases, 11 underwent the intraprocedural exclusionof the skull area areas with poor conditions, and the total number of sonications performed using this method was 30. Patient characteristics are shown in Table 1. Compared with the entire cohort, the SDR was significantly smaller in the cohort treated after excluding skull areas with poor conditions.

**Table 1.**
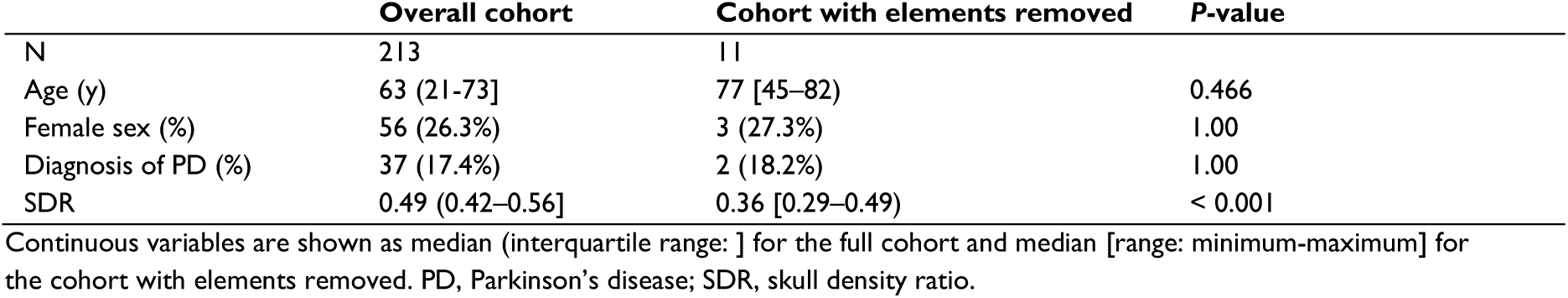
Patient characteristics.

|  | Overall cohort | Cohort with elements removed | P-value |
| --- | --- | --- | --- |
| N | 213 | 11 |  |
| Age (y) | 63 (21-73] | 77 [45-82) | 0.466 |
| Female sex (%) | 56 (26.3%) | 3 (27.3%) | 1.00 |
| Diagnosis of PD (%) | 37 (17.4%) | 2 (18.2%) | 1.00 |
| SDR | 0.49 (0.42-0.56] | 0.36 [0.29-0.49) | < 0.001 |

Continuous variables are shown as median (interquartile range: ] for the full cohort and median [range: minimum-maximum] for the cohort with elements removed. PD, Parkinson's disease; SDR, skull density ratio.

The frequency distributions of heating efficiency and the number of sonication elements are shown in Fig. 2. The median heating efficiency (°C/kJ) was 1.29 (minimum 0.027, first quartile 0.744, third quartile 1.97, maximum 5.67). The median number of sonication elements was 947 (minimum 719, first quartile 920, third quartile 966, maximum 1005). The median number of elements in cases where the unfavorable sonication fields were excluded was 962.5 (maximum 988, minimum 921) before application and 927.5 (maximum 961, minimum 865) after application, and the median number of excluded elements was 37.5 (maximum 74, minimum 6). We also visualized the relationship between sonication energy and temperature rise in cases that required more than 35,000 J of energy (i.e., cases that required considerable energy to raise the temperature and cases that were difficult to treat), among the most recent 100 cases, including cases where sonication areas with poor conditions were excluded (Fig. 3).

**Figure 2.**
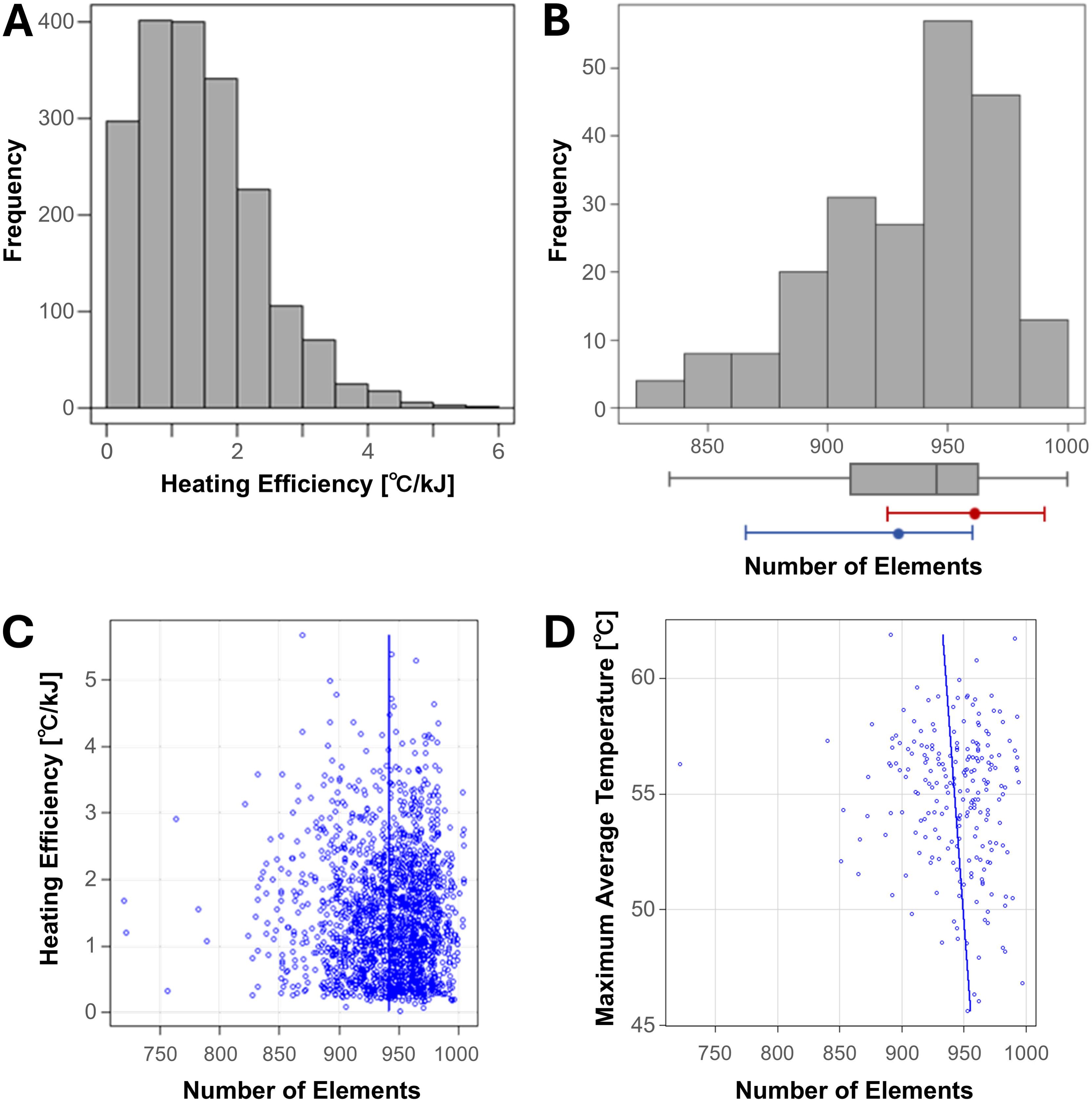
Statistical characteristics of heating efficiency and the number of sonication elements. (**A**) Frequency distribution of heating efficiency. (**B**) Frequency distribution of the number of sonication elements for all treatments. The blue and red bars represent cases where sonication areas with poor conditions were excluded (blue, before exclusion; red, after exclusion). The relationship between the number of sonication elements and the heating efficiency (**C**) and the maximum average temperature reached (**D**).

**Figure 3.**
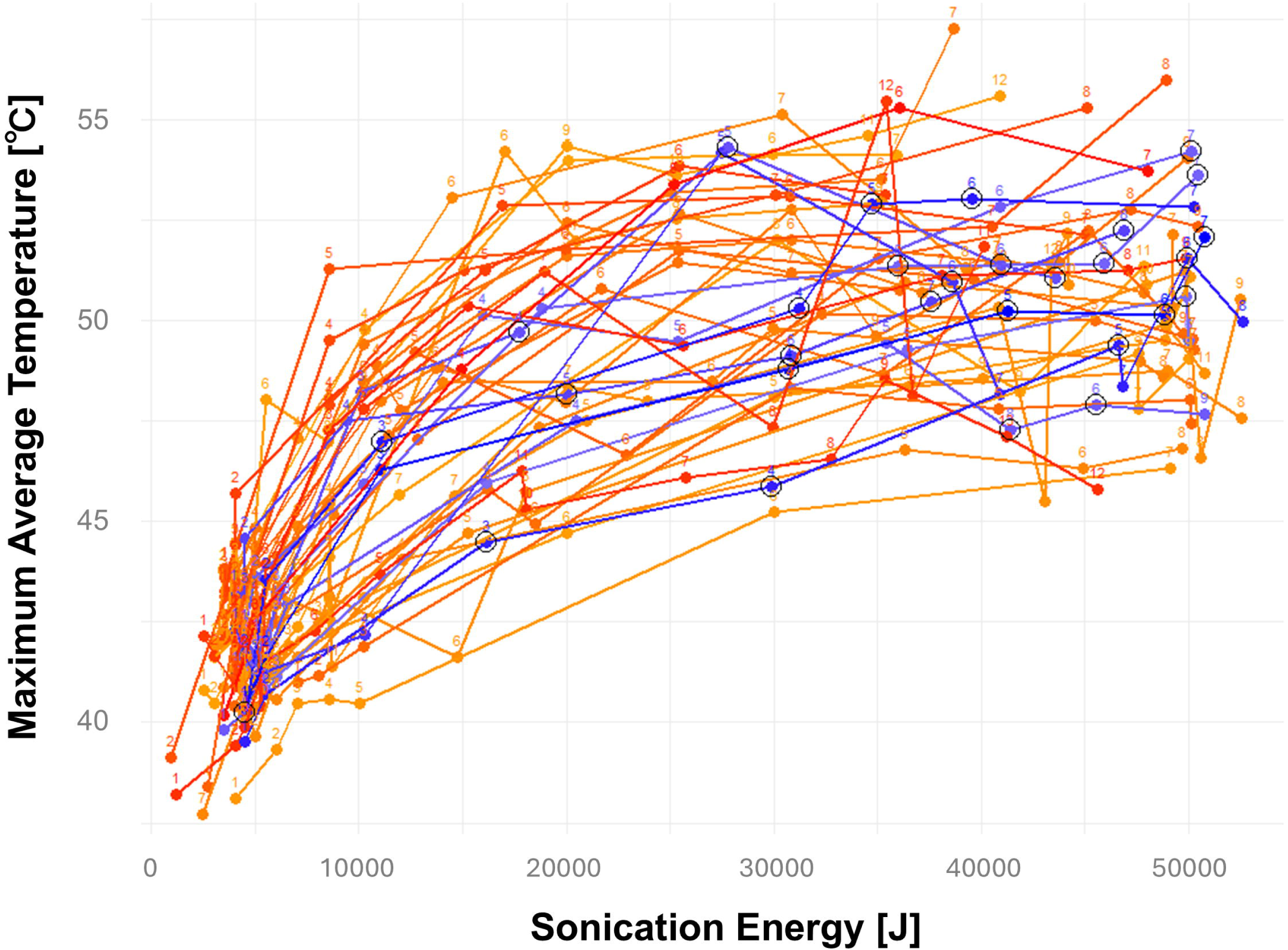
Relationship between the energy of each sonication and the temperature rise in cases where at least 35,000 J of energy was used. The sonications for the same case are connected by lines. The warm colors indicate cases treated using the regular method, and the cool colors indicate cases where the temperature rise was small after treatment began, and sonication areas with poor conditions were excluded. The sonications with the poor-conditioned area excluded are circled. Subscript denotes the sonication number.

The linear regression model for predicting heating efficiency, shown in Table 2, exhibited good accuracy (adjusted *R*^2^ = 0.769). Exclusion of the poor sonication field was a statistically significant predictor of heating efficiency (*P* = 0.0125). The regression coefficient had a positive value and indicated that exclusion of the poor sonication field increased the heating efficiency by 0.286 °C/kJ. The SDR, power, energy, and cumulative sonication count were also statistically significant predictors of heating efficiency (*P* = 1.15E^-12^, *P* = 1.38E^-19^, *P* = 1.03E^-4^, and *P* = 4.93E^-2^, respectively). There was no significance in the number of elements.

**Table 2.** Multiple regression model for predicting heating efficiency.

|  | Regression coefficient | Standard error | Confidence interval | P-value |
| --- | --- | --- | --- | --- |
| Intercept | 5.97E<sup>-1</sup> | 3.91E<sup>-1</sup> | -1.76E<sup>-4</sup> 1.37E<sup>-3</sup> | 1.29E<sup>-1</sup> |
| **Patient factors** |  |  |  |  |
| Age | -4.34E<sup>-3</sup> | 3.14E<sup>-3</sup> | -1.07E<sup>-5</sup> 2.00E<sup>-6</sup> | 1.74E<sup>-1</sup> |
| Sex (female) | 5.08E<sup>-2</sup> | 7.83E<sup>-2</sup> | -1.05E<sup>-4</sup> 2.07E<sup>-4</sup> | 5.19E<sup>-1</sup> |
| Diagnosis (PD) | 8.00E<sup>-2</sup> | 6.73E<sup>-2</sup> | -5.53E<sup>-5</sup> 2.15E<sup>-4</sup> | 2.40E<sup>-1</sup> |
| SDR | 5.23 | 4.08E<sup>-1</sup> | 4.42E<sup>-3</sup> 6.04E<sup>-3</sup> | 1.15E<sup>-22*</sup> |
| **Treatment factors** |  |  |  |  |
| Sonication power (W) | -1.25E<sup>-3</sup> | 1.17E<sup>-4</sup> | -1.48E<sup>-6</sup> -1.02E<sup>-6</sup> | 1.38E<sup>-19*</sup> |
| Sonication energy (J) | -1.13E<sup>-5</sup> | 2.80E<sup>-6</sup> | -1.69E<sup>-8</sup> -5.78E<sup>-9</sup> | 1.03E<sup>-4*</sup> |
| Sonication element number | 8.97E<sup>-5</sup> | 3.17E<sup>-4</sup> | -5.38E<sup>-7</sup> 7.18E<sup>-7</sup> | 7.78E<sup>-1</sup> |
| Cumulative sonication number | -9.44E<sup>-2</sup> | 3.71E<sup>-2</sup> | -1.89E<sup>-4</sup> -4.29E<sup>-7</sup> | 4.93E<sup>-2*</sup> |
| Cumulative temperature rise (°C) | 4.25E<sup>-3</sup> | 2.22E<sup>-3</sup> | -2.52E<sup>-7</sup> 8.75E<sup>-6</sup> | 6.36E<sup>-2</sup> |
| Exclusion of areas with poor conditions | 2.86E<sup>-1</sup> | 8.97E<sup>-2</sup> | 8.00E<sup>-5</sup> 4.92E<sup>-4</sup> | 1.25E<sup>-2*</sup> |

Adjusted R<sup>2</sup> = 0.769. \*P < 0.05. PD, Parkinson's disease; SDR, skull density ratio.

The adverse events are shown in Table 3. We did not observe any increase in the number of adverse events with the application of the method excluding the poor sonication field. The coagulation foci are shown in Fig. 4. We did not observe any difficulty in controlling the size of the coagulation foci with the application of the method excluding the poor sonication field.

**Figure 4.**
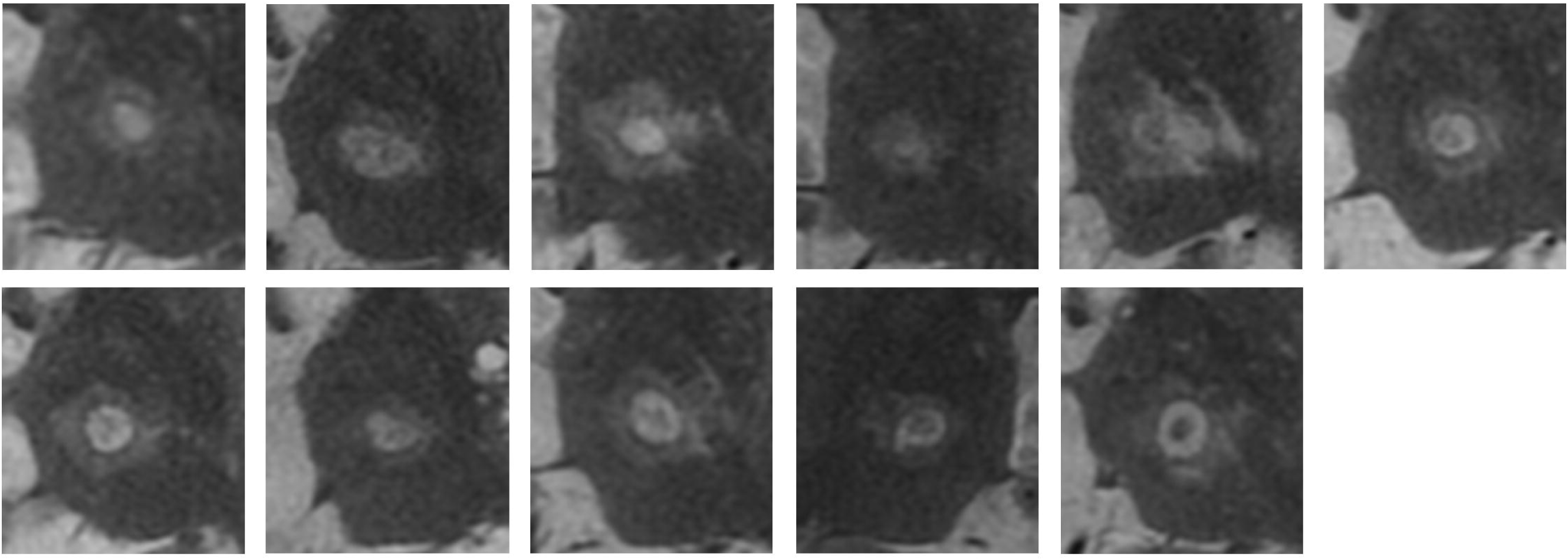
Coagulation foci in 11 cases where areas with poor conditions were excluded. T2-weighted image acquired the day after treatment.

**Table 3.** Adverse events after magnetic resonance-guided focused ultrasound surgery.

|  | Day after treatment | 3 months after treatment | 6 months after treatment |
| --- | --- | --- | --- |
| Headache | 2 | 0 | 0 |
| Nausea and vomiting | 1 | 0 | 0 |
| Dizziness | 1 | 0 | 0 |
| Scalp edema | 7 | 0 | 0 |
| Paresthesia and numbness | 4 | 1 | 1* |
| Dysarthria | 2 | 0 | 0 |
| Cerebellar ataxia | 1 | 0 | 0 |
| Motor weakness | 2 | 0 | 0 |
| Gait disturbance | 1 | 0 | 0 |
| Required inpatient rehabilitation | 1 | 0 | 0 |

\*Very mild. Data are numbers of adverse events.

### Representative case

A woman in her 70’s with a 10-year history of drug-resistant essential tremor and an SDR of 0.32 presented with a Clinical Rating Scale of Tremor score of 31. After thorough explanations and counseling, it was decided that thalamic coagulation using MRgFUS would be conducted. The sonication performed to verify efficacy and safety reached a maximum average temperature of 46°C at 850 W and 16,000 J. The temperature increased to 49°C at 1,100 W and 36,000 J, and then to 50°C at 1,200 W and 50,200 J (Fig. 5). We considered that the temperature rise necessary for target coagulation would be difficult to achieve with these settings and therefore excluded 48 elements from the sonication area (elements 928–970). The next sonication achieved a temperature rise of up to 51°C at 1,200 W and 50,200 J. The symptoms improved (Clinical Rating Scale of Tremor score = 9), and coagulation foci were also observed on the T2-weighted image.

**Figure 5.**
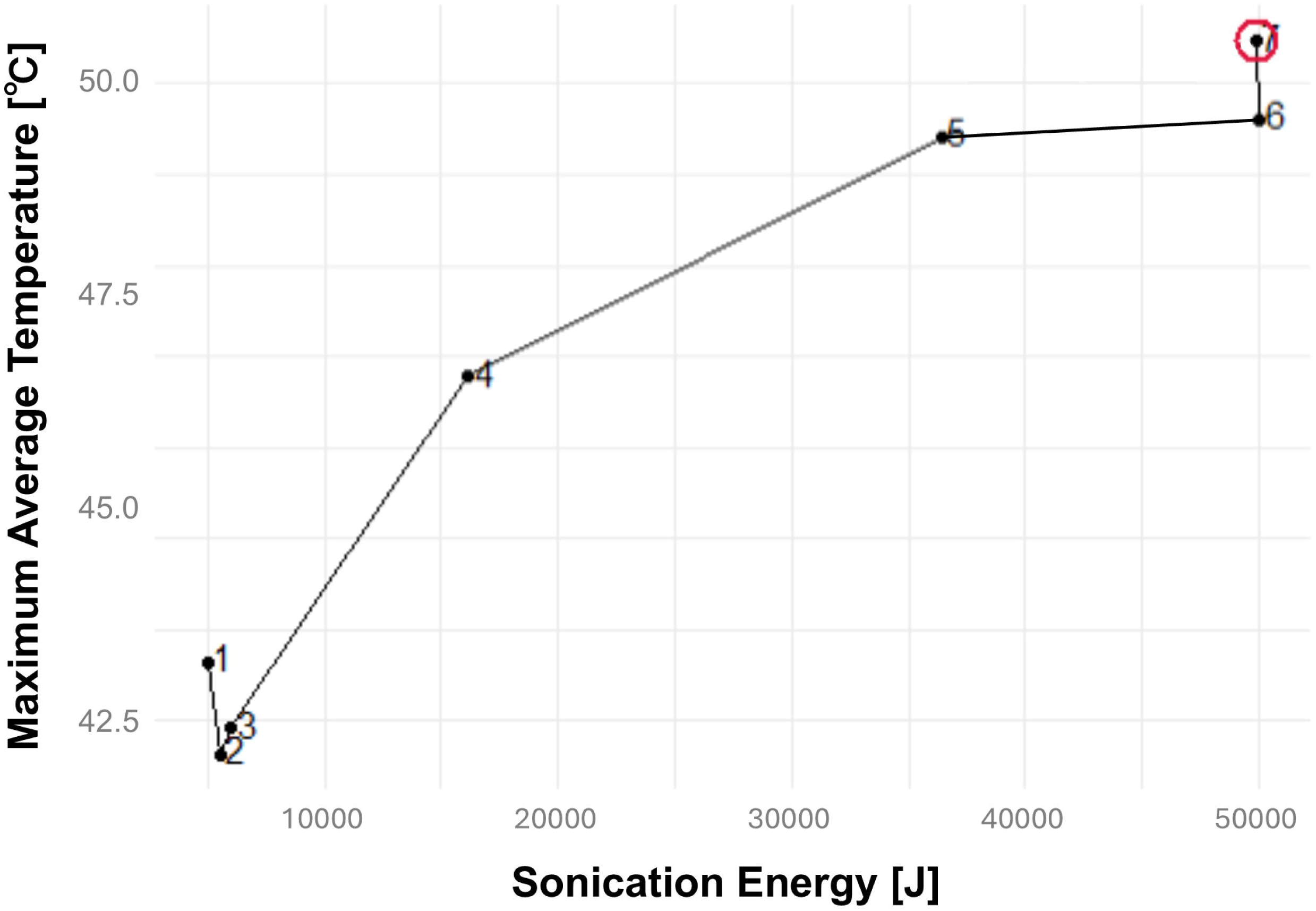
Representative case. A woman in her 70’s with essential tremor and a skull density ratio of 0.32. Left: Relationship between sonication energy and temperature rise. The red circle indicates sonication excluding areas with poor conditions. Subscript indicates the sonication number. Right: T2-weighted image of coagulation foci acquired the day after treatment.

## Discussion

In this study, it was found that the heating efficiency of MRgFUS could be improved by avoiding areas with a low SDR and thick skull. In the linear regression model, the regression coefficient for this application was 0.286. This result means that the heating efficiency was improved by 0.286°C/kJ using the method excluding the poor sonication field. If simply calculated, for every additional 10,000 J of isonication energy, the temperature increased by 2.86°C, which is a significant and important effect in clinical contexts: it may be possible to successfully treat cases that have proven difficult to treat so far.

Furthermore, the negative impact of reducing the total number of sonication elements was minimal. Although not statistically significant (*P* = 0.778), the regression coefficient for the number of sonication elements was 8.97E^-5^. This translates to an effect of 0.00897°C/kJ per 100 elements; in other words, when 100 elements are excluded, the temperature rise per 10,000 J of sonication energy is 0.0897°C smaller. This clinical effect is sufficiently smaller than the effect of excluding poorly conditioned areas to allow the negative impact on heating efficiency to be ignored.

Previous results of the simulation showed that when the number of elements decreases below 700, the coagulation clot size becomes difficult to control, which became one of the bases for the treatment protocol (Insightec Japan, personal communication, January 2025). In our results, the exclusion of poorly conditioned elements was carried out within the range where the number of elements did not fall below 850, and there was no clear increase in the difficulty of controlling the coagulation foci size or adverse events.

The results of this study also prompt two further suggestions regarding MRgFUS treatment approaches. First, reducing the number of sonications is expected to be effective;^12,16^ there has been one report of positive results with this method^18^, and the results of our study also support its application. The regression coefficient for cumulative sonication sessions was −0.0944 in this study, and a simple calculation indicates that for every additional sonication session, the temperature rise decreases by 0.0944°C. Second, as stated in a previous report^17^, sonication power had a negative effect on heating efficiency. The regression coefficient was −0.00125, and a simple calculation indicates that for every 100 W increase in sonication power, the temperature rise per 10,000 J of energy will be reduced by 1.25°C. On this basis, if the same amount of energy is delivered, it may be beneficial to use less power to achieve a greater temperature rise.

Our study has several limitations. First, it is a retrospective study. Second, no specific selection criteria for the cases were used. Third, the criteria for excluding elements from the sonication field were not consistent. Fourth, the number of elements excluded from the sonication field was not specified. Therefore, verification of the results by a prospective study is required.

In conclusion, we found that the heating efficiency of MRgFUS was significantly improved when sonication was conducted while avoiding areas with a low SDR and thick skull. Using this approach, it may be possible to successfully treat cases that have proven difficult to treat so far.

## Data availability

The datasets generated and/or analyzed during the current study are available in the Dryad repository, accessible via https://doi.org/10.5061/dryad.qjq2bvqs7. []

## Acknowledgements

We thank Michael Irvine, PhD, from Edanz (https://jp.edanz.com/ac) for editing a draft of this manuscript.

## Previous Presentations

The 20^th^ Biennial Meeting of the World Society for Stereotactic & Functional Neurosurgery, 2024/09/04, Chicago, USA, e-Poster Presentation.

